# A Deep Learning Approach for Automated Extraction of Functional Status and New York Heart Association Class for Heart Failure Patients During Clinical Encounters

**DOI:** 10.1101/2024.03.30.24305095

**Authors:** Philip Adejumo, Phyllis Thangaraj, Lovedeep Singh Dhingra, Arya Aminorroaya, Xinyu Zhou, Cynthia Brandt, Hua Xu, Harlan M Krumholz, Rohan Khera

**Affiliations:** Section of Cardiovascular Medicine, Department of Internal Medicine, Yale School of Medicine, New Haven, CT; VA Connecticut Healthcare System, West Haven, CT, USA; Section of Health Informatics, Department of Biostatistics, Yale School of Public Health, New Haven, CT; Biomedical Informatics and Data Science, Yale School of Medicine, New Haven, CT; Department of Health Policy and Management, Yale School of Public Health, New Haven, CT, USA; Center for Outcomes Research and Evaluation, Yale-New Haven Hospital, New Haven, CT

## Abstract

**Introduction:** Serial functional status assessments are critical to heart failure (HF) management but are often described narratively in documentation, limiting their use in quality improvement or patient selection for clinical trials. We developed and validated a deep learning-based natural language processing (NLP) strategy to extract functional status assessments from unstructured clinical notes.

**Methods:** We identified 26,577 HF patients across outpatient services at Yale New Haven Hospital (YNHH), Greenwich Hospital (GH), and Northeast Medical Group (NMG) (mean age 76.1 years; 52.0% women). We used expert annotated notes from YNHH for model development/internal testing and from GH and NMG for external validation. The primary outcomes were NLP models to detect (a) explicit New York Heart Association (NYHA) classification, (b) HF symptoms during activity or rest, and (c) functional status assessment frequency.

**Results:** Among 3,000 expert-annotated notes, 13.6% mentioned NYHA class, and 26.5% described HF symptoms. The model to detect NYHA classes achieved a class-weighted AUROC of 0.99 (95% CI: 0.98-1.00) at YNHH, 0.98 (0.96-1.00) at NMG, and 0.98 (0.92-1.00) at GH. The activity-related HF symptom model achieved an AUROC of 0.94 (0.89-0.98) at YNHH, 0.94 (0.91-0.97) at NMG, and 0.95 (0.92-0.99) at GH. Deploying the NYHA model among 166,655 unannotated notes from YNHH identified 21,528 (12.9%) with NYHA mentions and 17,642 encounters (10.5%) classifiable into functional status groups based on activity-related symptoms.

**Conclusions:** We developed and validated an NLP approach to extract NYHA classification and activity-related HF symptoms from clinical notes, enhancing the ability to track optimal care and identify trial-eligible patients.

## BACKGROUND

Heart failure (HF) is characterized by a spectrum of symptoms that often impair patients’ functional ability and adversely affect their quality of life.^1,2^ Clinical practice guidelines recommend regular functional status evaluations using the New York Heart Association (NYHA) classification system, which grades HF severity based on physical activity limitations and symptoms.^3,4^ These assessments are crucial for informing therapeutic decisions, such as selecting appropriate guideline-directed medical therapies and determining the need for primary prevention implantable cardioverter defibrillators (ICDs).^5^

While regular functional status assessments are expected to be a standard component of clinical management for patients with HF, these assessments are primarily documented in unstructured medical notes.^6–8^ The consistency with which they guide care has only been evaluated in limited manual abstraction studies of clinical documentation.^9^ The absence of standardized systems to capture functional classes and variations in clinical documentation practices hinder evaluating how well HF treatment decisions align with clinical guidelines across a wide range of patients. Furthermore, clinical trials are increasingly recruiting directly from electronic health records (EHRs), but the inability to identify a patient’s functional status in a computable manner restricts this approach.

To address this, we developed and validated efficient, lightweight, transformer-based deep learning models for natural language processing (NLP) to automate the extraction of recorded functional status assessments within unstructured clinical notes.^10^ The objective of this study is to develop a scalable, AI-driven approach that can reliably identify both explicit mentions of NYHA classes and categorize relevant functional status descriptors within the vast array of unstructured text in electronic health records (EHRs).^11^

## METHODS

The Yale Institutional Review Board reviewed the study, which approved the protocol and waived the need for informed consent, as it represents a secondary analysis of existing data.

### Data Source

We used data from the Yale New Haven Health System (YNHHS) EHR from 2013 to 2022, encompassing an academic hospital and a multisite community practice network with seven distinct sites. YNHHS caters to a diverse demographic, and the community it serves reflects the national US population across age, sex, race, ethnicity, and socioeconomic status.^12^ We extracted key structured and unstructured EHR data for the study population from Yale-New Haven Health System’s Epic® Clarity database. The structured fields included patient demographics, diagnosis, procedure codes, and healthcare encounters. The unstructured data included medical notes, which provide a comprehensive clinical history for each patient.

### Study Population

The study cohort comprised patients diagnosed with HF who had at least one healthcare encounter within any of the YNHHS Heart and Vascular Center Outpatient Clinics —including Yale New Haven Hospital (YNHH), Northeast Medical Group (NMG), and Greenwich Hospital (GH)—between January 1, 2013, and June 30, 2022 (**Supplemental Figure 1, Supplemental Table 1**). Eligible patients were 18 years or older and had one or more healthcare encounters with an International Classification of Diseases Ninth and Tenth Editions (ICD 9/10) code for HF. The definition prioritized sensitivity over specificity of individuals who might have HF and being evaluated in ambulatory practices. We also identified comorbidities, including acute myocardial infarction (AMI), cardiomyopathy, hypertension (HTN), diabetes, and chronic kidney disease (CKD), using the relevant ICD-9 and ICD-10 diagnosis codes (**Supplemental Table 2**). Medical documentation, including history and physicals, progress notes, letters, and assessments and plans, were examined at the individual encounter level to provide a comprehensive view of the clinical journey (**Figure 1**). This method allowed for the inclusion of diverse documented information, capturing various facets of each encounter that could reflect the patient’s health status, treatment adjustments, and functional status as recorded by different healthcare providers across multiple visits.

**Figure 1.**
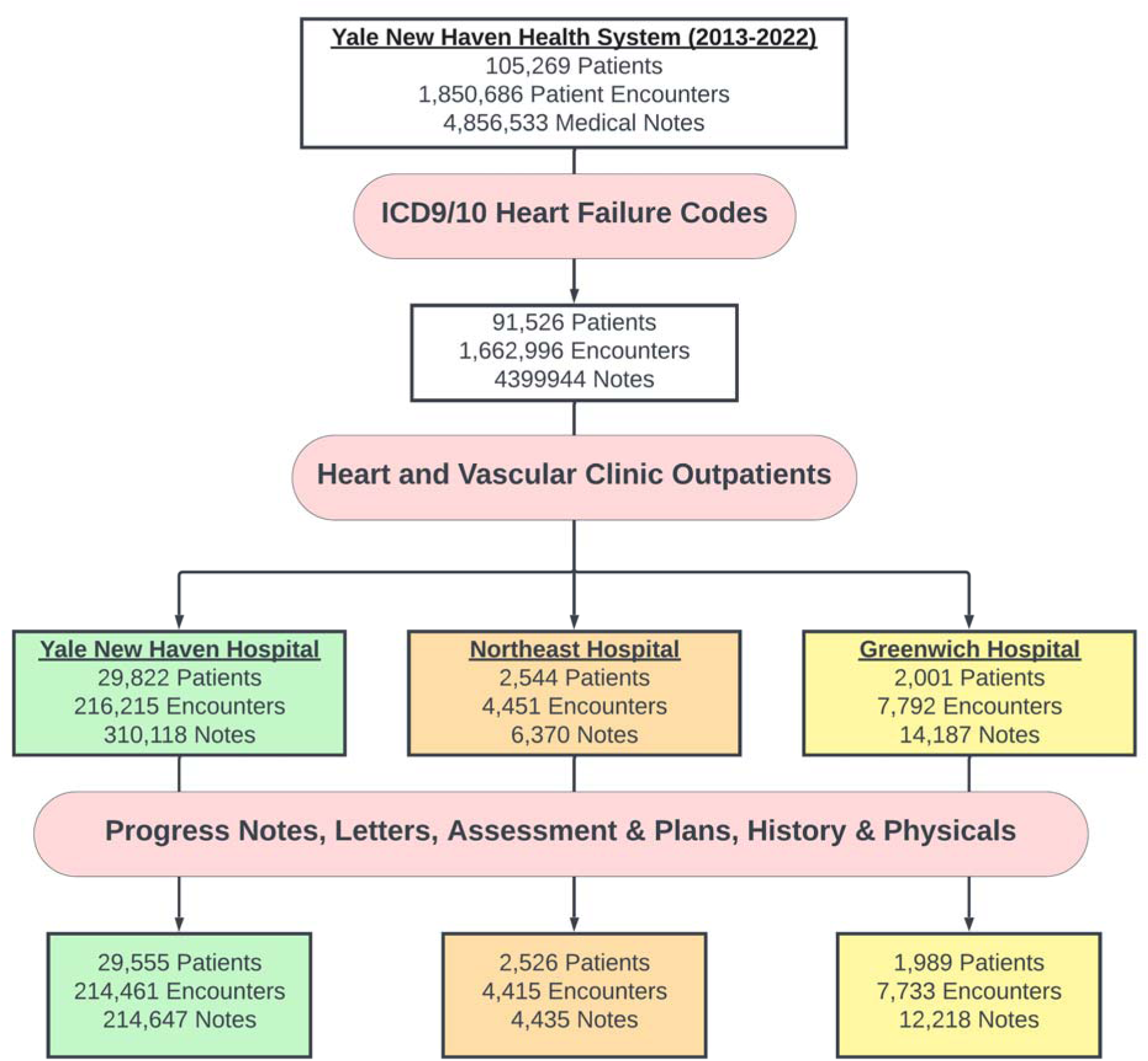
Development of Study Cohort Flowchart. The strategy employed to develop the heart and vascular clinical outpatients’ cohort at Yale New Haven Health System and gather the clinical documentation. Abbreviations: ICD, International Classification of Disease.

### Study Outcomes

We evaluated a series of outcomes, including (a) the performance of our deep learning models in identifying explicit mentions of NYHA classes in unstructured medical notes against expert chart abstraction, (b) the performance of a model designed to extract activity/rest-related HF symptoms based on expert annotation of the notes, and (c) a descriptive evaluation of functional status assessment frequency across all outpatient encounters.

### Data Pre-processing

We sectioned all notes to include only the history of presenting illness or the assessment and plan. This focused the analysis on the most relevant sections for assessing functional status and NYHA classification. We leveraged MedSpaCy’s clinical sectionizer to tag and remove other sections of the outpatient medical note, including the past medical history, labs, studies, allergies, physical exam, medications, family history, and imaging.^13^

In the selected components of the note, we pursued a sentence-level annotation process to identify sentences containing information relevant to the patient’s functional status and NYHA classification. For this, we split the notes into individual sentences using a clinically trained transformer-based sentence tokenizer. Each sentence was then labeled as either containing functional status information or not. The labeled sentences were later reassembled to reconstruct the context of each note, enabling note-level classification based on the aggregated sentence-level labels. This step was critical for ensuring the integrity of the note-level classification.

### Manual Annotation

Annotators labeled 2,000 randomly selected outpatient notes at YNHH at the note level for the specified document classification labels. In addition, we annotated a separate set of 1,000 notes from outpatient clinics at GH and NMG for additional external validation (500 each) **(Figure 2)**. Specific details of the annotating schema for the NLP document classification models can be found in the annotation guidelines **(Annotating Guidelines in Supplement)**. The annotation focused on labeling each note for the explicit mention of NYHA classification and the presence of HF symptoms associated with activity or rest **(Table 2)**. To guide the annotation, three expert annotators collaboratively established an annotation scheme that delineated the criteria for class identification and symptom extraction at the note level. Each note was evaluated for relevant features based on its constituent sentences, and these features were aggregated to inform the overall annotation of the entire note, ensuring a standardized and precise approach across the dataset.

**Figure 2.**
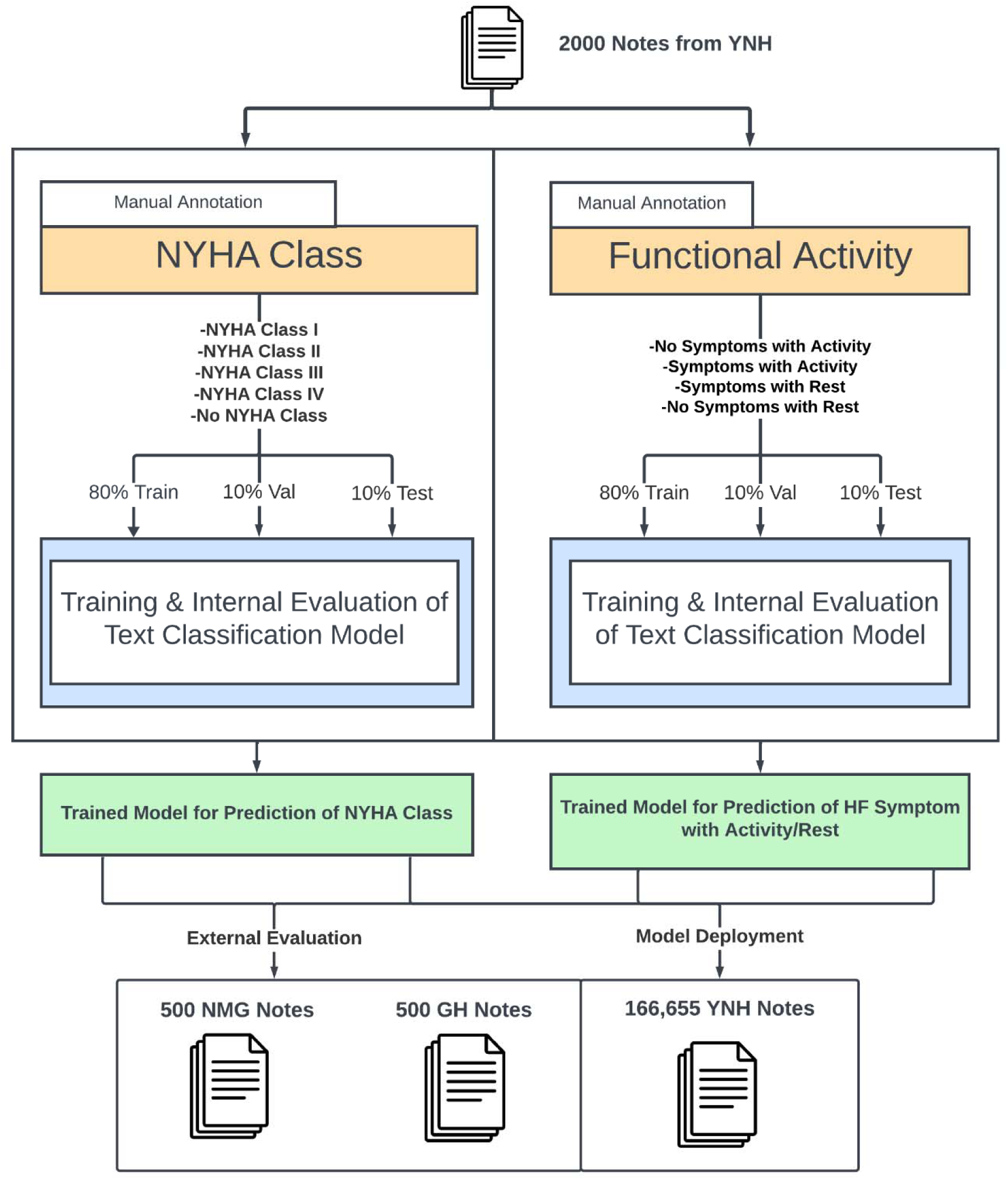
Annotating Scheme/Study Overview. The overview of the study, including (1) the annotation scheme established and employed to annotate New York Heart Association class and functional activity to the clinical documentations at Yale New Haven Health System, and (2) the model development, internal and external validation, and deployment strategies. Abbreviations: GH, Greenwich Hospital; NMG, Northeast Medical Group; NYHA, New York Heart Association; YNH, Yale New Haven Hospital.

### Model Development

We randomly separated the annotated dataset from YH into three subsets: 80% was allocated for training the models, 10% for validation to fine-tune the hyperparameters, and the remaining 10% comprised unseen notes dedicated to internal testing. We chose to develop a ClinicalBERT-based model, a lightweight state-of-the-art variant of the Bidirectional Encoder Representations from Transformers (BERT) architecture. ClinicalBERT has been specifically pre-trained on a large corpus of clinical text, making it well-suited for capturing the nuances and terminology present in medical notes, with ability to fine-tune for specific applications.^14^ The fine-tuning process involved adjusting the ClinicalBERT model weight parameters to better capture the nuances and terminology specific to our two independent document classification tasks: Distinguishing between the various NYHA functional classifications (Class I, Class II, Class III, Class IV, or no mention of NYHA class) and identifying the documented symptoms of HF occurring during activity (symptoms with activity, lack of symptoms with activity, or no mention) or at rest (symptoms with rest, lack of symptoms with rest, or no mention).

### Interpretability Analysis

For our interpretability analysis, we applied an adapted version of the SHapley Additive exPlanations (SHAP) method specifically designed for transformer-based classifiers like BERT. This adapted SHAP method quantifies the impact of each feature on the NLP models’ outputs while accounting for the complex feature interactions in transformer architectures.^15,16^ We selected a stratified sample of 100 random notes from each NYHA class to ensure that the analysis would reflect a broad spectrum of clinical scenarios. Using the SHAP Explainer, we permuted features within these notes to create 2,000 synthetic examples, introducing a diverse array of text lengths and complexities into the analysis. The resulting SHAP values for these permutations provided insight into the significance of individual words and phrases in the notes. We calculated and aggregated these SHAP values to assess the mean positive impact of each feature, enabling us to determine the most influential factors in our model’s decision-making process.

### Model Defined Prevalence of Functional Status Assessments

To demonstrate the clinical utility of our models, we evaluated the frequency of NYHA class documentation and discussed activity-related symptoms in the remaining large EHR dataset from YNHHS that was not used in model development or validation. These encounters were similar to those with one or more HF encounters but without explicit manual annotation of these encounters. We applied the NYHA class model to identify explicit mentions of NYHA classes in these records and used the HF symptom association model to classify additional records based on the presence of activity-related symptoms. Furthermore, we employed the HF symptom association model to re-categorize patients without explicit mentions of NYHA class in their clinical documentation to different NYHA classes (**Supplemental Figure 2**). By combining the explicit mentions identified by the NYHA class model and the additional classifications provided by the symptom association model, we sought to enhance the capture of functional status information and provide a more comprehensive assessment of HF severity across the patient population.

To further illustrate the potential impact of our models, we assessed the frequency of NYHA class documentation in the year preceding the implantation of an ICD, a key HF management decision that requires assessment of NYHA Class in making the determination of eligibility in clinical practice guidelines.^17^ We identified patients who underwent ICD implantation using the relevant procedure codes and determined the presence of explicit NYHA class mentions in their clinical notes the year before the procedure (**Supplemental Table 3**). This analysis aimed to showcase the utility of our models in identifying clinically relevant information during critical windows of care, such as the period leading up to a major procedure like ICD implantation.

### Statistical Analysis

For all descriptive analyses, categorical variables were reported as frequency and percentage, while continuous variables were summarized as mean and standard deviation. Given the multiclass nature of our analysis, we assessed their performance using both micro-averaged and macro-averaged metrics. Micro-averaged metrics were chosen as the primary focus of our analysis, as they provide a comprehensive assessment of the model’s performance across individual classes, giving equal weight to each classification decision. This approach is particularly suitable for our study, as it aligns with our objective of evaluating the overall performance of the NLP models in identifying functional status information across the entire dataset. Macro-averaged metrics, which treat all classes equally, regardless of their frequency, were also reported to provide additional insights into the model’s performance on individual classes. The metrics incorporated in the analysis include the area under the receiver operating characteristic curve (AUROC), areas under the precision-recall curves (AUPRC), accuracy, precision (or positive predictive value), recall (or sensitivity), specificity, and F_1_-score. The models produced continuous probabilities, which we dichotomized for the presence or absence of the condition by thresholds that maximized Youden’s Index, ensuring a balance between sensitivity and specificity. For each of these metrics, 95% confidence intervals were obtained from bootstrap resampling with 1,000 iterations. A Python-based computational environment was used for all analysis. Libraries consisted of Pandas (1.5.3) for data processing, Numpy (1.23.5) and SciPy (1.10.1) for numerical operations, and Scikit-learn (1.2.2) for machine learning tasks. Corpus annotation was conducted using Prodigy (1.12), and model development and refinement using SpaCy (3.6).

## RESULTS

### Study Cohort

There were 34,070 patients who had at least one healthcare encounter at a cardiovascular outpatient clinic within YNHHS between January 1, 2013 and June 30, 2022, and had a recorded diagnosis of HF. This dataset encompassed records from 29,555 patients at YNHH (214,461 encounters; 214,647 clinical notes), 2,526 patients at NMG (4,415 encounters; 4,435 notes), and 1,989 patients at GH (7,733 encounters; 12,218 notes). The average age of the cohort was 76.1 years (±12.6), with 17,728 women (52.0%). 47,109 (77.9%) of the patients in the cohort were White (77.9%), and 4,422 (14.2%) were Black. Among these, 27,437 (71.3%) patients had a diagnosis of hypertension, 14,580 (38.7%) had diabetes, and 10,648 (28.5%) had chronic kidney disease, with other clinical characteristics of the cohorts reported in **Table 1**.

**Table 1.** Demographic Characteristics Across Study Sites. This table delineates patient demographics, note types, and comorbidities across Yale-New Haven Hospital, Northeast Medical Group, and Greenwich Hospital, highlighting variations in age, sex, race, ethnicity, and health conditions.

| Characteristic |  | Yale-New Haven Hospital | Northeast Medical Group | Greenwich Hospital |
| --- | --- | --- | --- | --- |
| Age, year (Standard Deviation) |  | 76.57 (13.45) | 73.64 (12.85) | 79.07 (11.54) |
| Sex |  |  |  |  |
|  | Male (%) | 14,171 (47.96%) | 1,093 (43.28%) | 1,078 (54.20%) |
|  | Female (%) | 15,384 (52.04%) | 1,433 (56.72%) | 911 (45.80%) |
| Race |  |  |  |  |
|  | American Indian or Alaska native (%) | 93 (0.32%) | 12 (0.05%) | 0 (0.00%) |
|  | Asian (%) | 345 (1.18%) | 35 (0.14%) | 54 (1.87%) |
|  | Black (%) | 4,245 (14.50%) | 106 (0.43%) | 71 (2.45%) |
|  | Native Hawaiian or Other Pacific Islander (%) | 48 (0.16%) | 2 (0.01%) | 7 (0.24%) |
|  | White (%) | 23,147 (79.12%) | 22,216 (91.07%) | 1,746 (60.29%) |
|  | Unknown (%) | 435 (1.49%) | 2,216 (9.09%) | 23 (0.79%) |
|  | Other (%) | 1,242 (4.24%) | 76 (0.31%) | 88 (3.04%) |
| Ethnicity |  |  |  |  |
|  | Non-Hispanic (%) | 1873 (6.33%) | 92 (3.64%) | 95 (4.77%) |
|  | Hispanic or Latino (%) | 27,295 (92.35%) | 2,338 (92.55%) | 1,876 (94.31%) |
|  | Unknown (%) | 387 (1.30%) | 96 (3.80%) | 18 (0.90%) |
| Demographics |  |  |  |  |
|  | Number of Patients | 29,822 | 2,544 | 2,001 |
|  | Number of Encounters | 216,215 | 4,451 | 7,792 |
| Comorbid Conditions |  |  |  |  |
|  | Acute Myocardial Infarction (%) | 4,435 (7.77%) | 288 (7.04%) | 222 (5.99%) |
|  | Cardiomyopathy (%) | 9,451 (16.55%) | 955 (23.35%) | 592 (15.99%) |
|  | Hypertension (%) | 25,106 (43.95%) | 2,159 (52.78%) | 1,729 (46.73%) |
|  | Diabetes (%) | 12,951 (22.69%) | 926 (22.64%) | 703 (18.99%) |
|  | Chronic Kidney Disease (%) | 9,530 (16.69%) | 662 (16.18%) | 456 (12.32%) |

**Table 2.** Definitions and Pertinent Examples of Functional Status Labels. This table presented the definitions of the New York Heart Association classes and Heart Failure symptoms analyzed using the two multiclass classification Natural Language Processing models developed in this study, along with pertinent deidentified examples. Abbreviations: HF, Heart Failure; NYHA, New York Heart Association.

| Label | Definition | Pertinent Example |
| --- | --- | --- |
| NYHA Class Model |  |  |
| NYHA Class I | A widely recognized system for categorizing heart failure severity based on symptoms and limitations in physical activity. It ranges from Class I (no symptoms) to Class IV (severe limitations). | “Patient presents with NYHA Class II symptoms, experiencing slight limitation during ordinary activity.” |
| NYHA Class II |  |  |
| NYHA Class III |  |  |
| NYHA Class IV |  |  |
| HF Symptom Association Model |  |  |
| HF Symptoms Associated with Activity | Descriptions of heart failure symptoms that become evident or worsen during physical exertion. | “During moderate exercise, the patient reports shortness of breath and palpitations.” |
| Lack of HF Symptoms Associated with Activity | Refers to the absence of heart failure symptoms during physical activity, indicating a lesser severity of HF. | “Patient shows no signs of fatigue or dyspnea during routine physical activities.” |
| HF Symptoms Associated with Rest | Descriptions of heart failure symptoms that persist or are present even when the patient is at rest. | “Patient experiences fatigue and dyspnea, even while at rest.” |
| Lack of HF Symptoms Associated with Rest | Indicates the absence of heart failure symptoms while the patient is at rest, suggesting a controlled or mild form of HF. | “Patient reports feeling comfortable and symptom-free while resting.” |

### Manual Annotation

Of the 2,000 clinical notes from YNHH that were expert annotated, 271 (13.6%) had any mention of NYHA class, with 57 (2.9%) Class I, 118 (5.9%) Class II, 86 (4.3%) Class III, and 10 (0.5%) Class IV **(Figure 3, Supplemental Table 4)**. The remaining did not mention an NYHA class. In the same set, activity-related symptoms of HF were reported in 486 (24.3%) of the notes, with 329 (16.5%) reporting the absence of symptoms with activity. Symptoms at rest were noted in 45 (2.3%) of cases **(Figure 3, Supplemental Table 5)**. Of the 500 expert annotated notes from NMG and GH, NYHA classes were explicitly noted in 80 (16.0%) and 23 (4.6%) of the notes, respectively **(Figure 3, Supplemental Table 4)**. In addition, activity-related HF symptoms were reported in 125 (25%) and 39 (7.8%) of the notes at NMG and GH, respectively. The absence of symptoms with activity was found in 54 (10.8%) and 38 (7.6%) of the notes at NMH and GH, respectively **(Figure 3, Supplemental Table 5)**.

**Figure 3.**
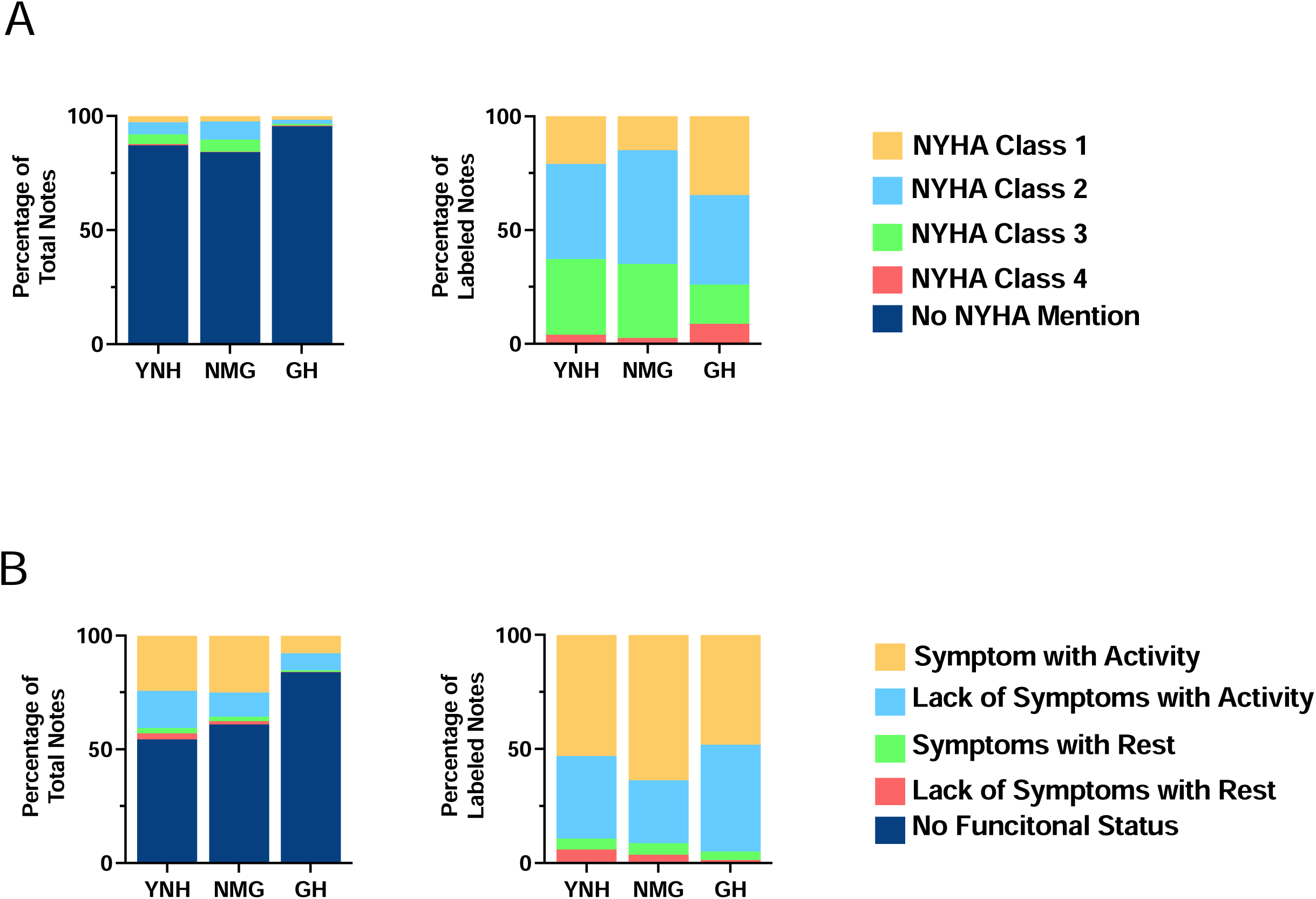
Distribution of Annotated Labels Annotation Stacked Bar Chart. The percentage of notes assigned to each category at each outpatient clinics of Yale New Haven Health System as manually annotated by experts. Abbreviations: GH, Greenwich Hospital; NMG, Northeast Medical Group; NYHA, New York Heart Association; YNH, Yale New Haven Hospital.

### Model Evaluation

The NYHA classification model demonstrated robust performance in identifying the presence of NYHA class documentation, with a micro-averaged AUROC of 0.99 (95% CI: 0.93-1.00) at YNHH, 0.98 (95% CI: 0.96-1.00) at NMG, and 0.98 (95% CI: 0.92-1.00) at GH **(Figure 4, Table 3)**. The corresponding AUPRCs ranged from 0.22 to 1.00. Similarly, the model performance for classifying individual NYHA classes ranged from 0.96 to 1.00 for classes I, II, III, and IV at YNHH **(Supplemental Table 6)**. The HF symptom association model, which identifies the presence of activity-related HF symptoms, displayed consistent performance across all sites, with micro-averaged AUROCs of 0.94 (95% CI: 0.89-0.98) at YNHH, 0.94 (95% CI: 0.91-0.97) at NMG, and 0.95 (95% CI: 0.92-0.99) at GH **(Figure 4, Table 3)**. The model performed well at detecting the presence (AUROC, 0.98; 95% CI: 0.96-0.99) and absence (0.99; 95% CI: 0.99-1.00) of HF symptoms with activity, as well as the presence (0.94; 95% CI: 0.92-0.96) and absence (0.92; 95% CI: 0.90-0.95) of symptoms at rest. The corresponding AUPRCs ranged from 0.66 to 0.98 (All metrics reported in **Supplemental Table 6)**.

**Figure 4.**
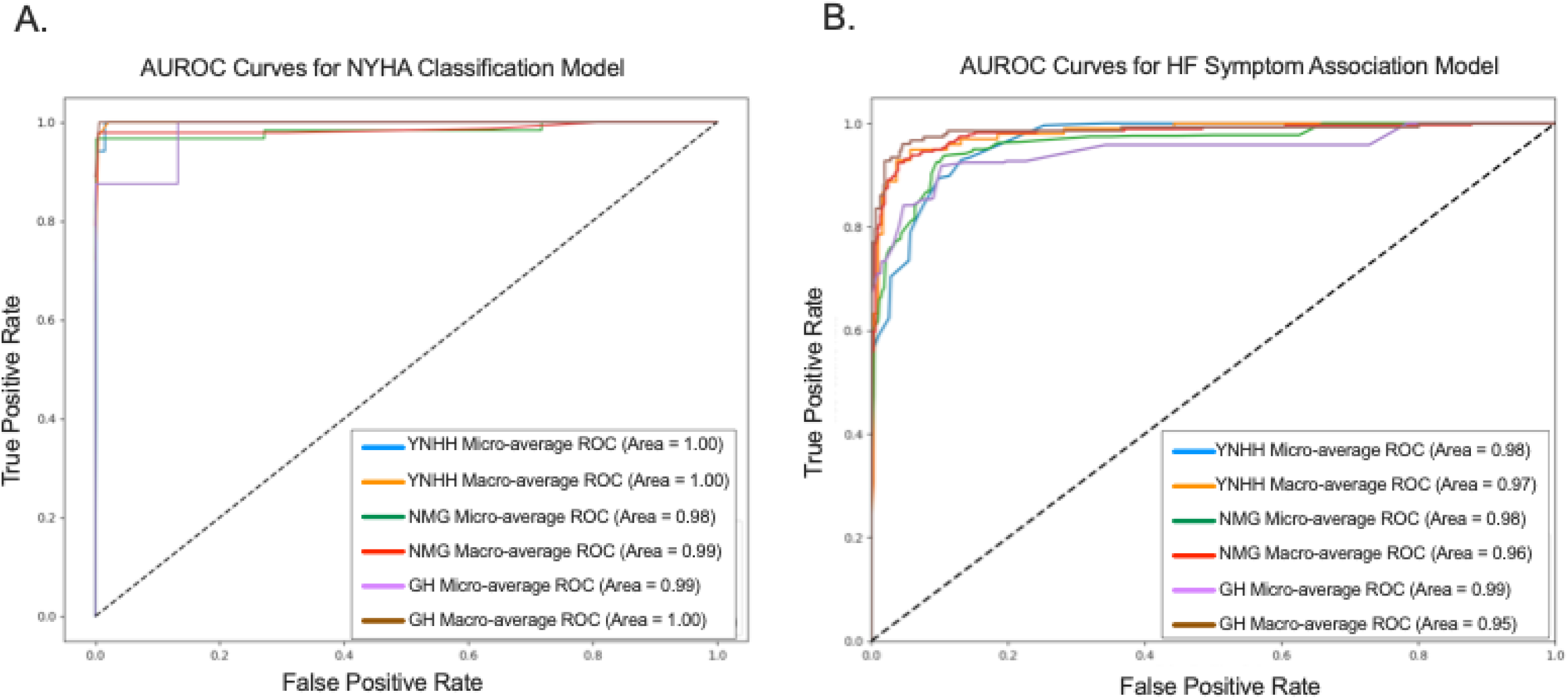
Micro and Macro Area Under the Receiver Operating Characteristic Curve of Natural Language Processing Models in Characterizing Functional Status Labels. Abbreviations: AUROC, area under the receiver operating characteristic curve; GH, Greenwich Hospital; NMG, Northeast Medical Group; NYHA, New York Heart Association; YNHH, Yale New Haven Hospital.

**Table 3.**
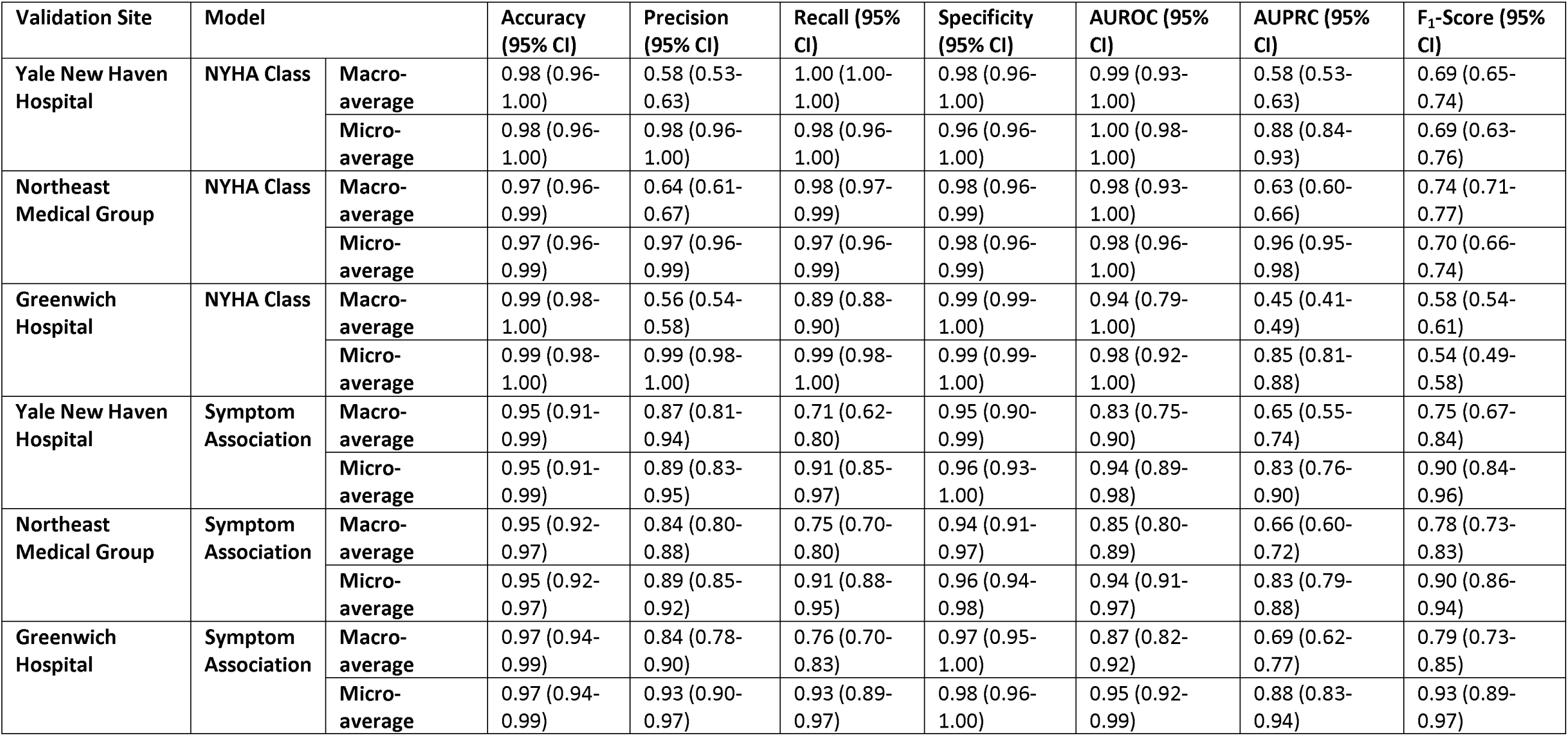
Performance Metrics of Natural Language Processing Models in Classifying Functional Status Labels. This table presents the micro average performance of the New York Heart Association class model and symptom association model, respectively, measured by accuracy, precision, recall, specificity, area under the receiver operating characteristic curve, area under the precision recall curve, and F_1_-score. 95% confidence intervals were obtained from bootstrap sampling with 1,000 iterations. Abbreviations: AUROC, area under the receiver operating characteristic curve; AUPRC, area under the precision recall curve; NYHA, New York Heart Association.

### Interpretability Analysis

The SHAP-based interpretability analysis revealed key features the NLP models leveraged for classifying the NYHA classes and the presence of HF symptoms **(Figure 5)**. In the model for NYHA classification, designations such as ’NYHA’, ’Class’, and the corresponding Roman numerals ’I’, ’II’, ’III’, and ’IV’ were the highest weighted features, consistent with the classification criteria detailed in our annotation guidelines. For the HF symptom association model, clinical descriptors of symptoms, particularly ’dyspnea’ and ‘breathlessness’ alongside verbs correlating with physical exertion or movement like ‘standing’ and ‘exertional’, were the highest weighted features. This delineation of terms highlights the models’ capacity to extract and prioritize clinical language reflective of patients’ functional status and HF severity from the unstructured text data.

**Figure 5.**
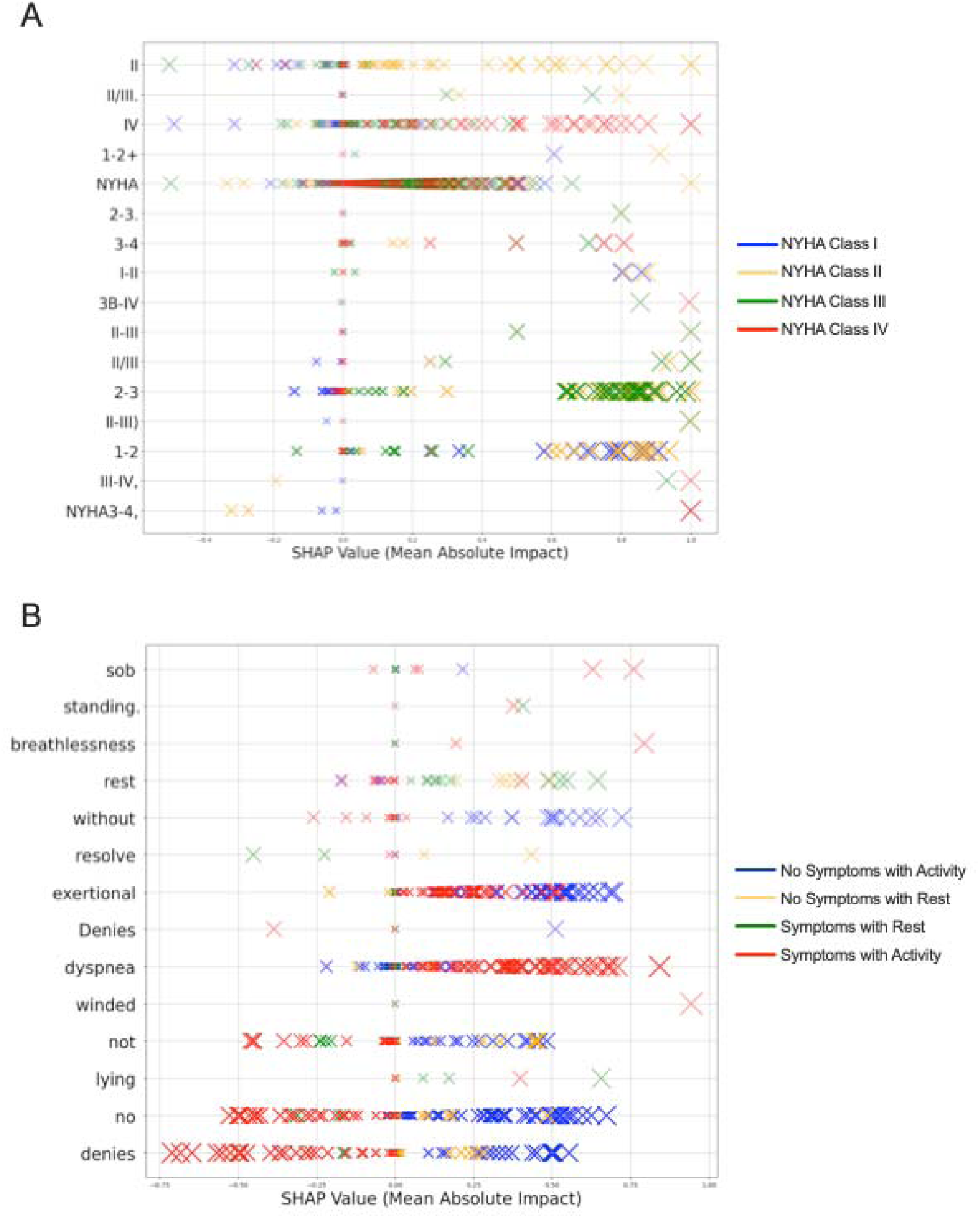
Top 15 Features by Mean Positive SHAP Value Across NYHA Class and HF Symptom Association Models. SHAP explainer plot based on stratified sample of 100 random notes from each NYHA class (400 notes in total), illustrating the impact of each of the top 15 features on model prediction. Each “X” represented a clinical note, with the absolute value in the x-axis (or the size of the “X”) representing the impact of the token presented on the y-axis on model prediction. Large size of “X” or larger absolute value on the x-axis represented larger impact on the model output. Colors represented model prediction of each clinical note. Abbreviations: NYHA, New York Heart Association.

### Evaluation of NYHA Classification and activity-related symptoms across notes

Across the documentation of all 166,655 outpatient encounters at YNHH not used in either model development or validation, our NYHA-extraction algorithm identified explicit mentions of NYHA classes in 21,528 (12.9%) encounters, or approximately 1 in 8 encounters. These were classified across the different classes, with 9,878 (5.9%) for Class I, 10,824 (6.5%) for Classes II or III, and 826 (0.5%) for Class IV **(Table 4, Figure 6)**. The model that extracted descriptive mentions of activity/rest-related HF symptoms could be used to additionally assign 17,642 (10.5%) encounters to an NYHA class, including 7,670 encounters as Class I (4.6%), 9,094 as Classes II or III (5.4%), and 788 as Class IV (0.5%) **(Table 4, Figure 6)**. Combining explicit mentions and NLP re-categorizations resulted in a functional status classification in 39,170 (23.5%) of all encounters, or nearly 1 in 4 encounters, representing an 82% increase in information capture compared to explicit mentions alone **(Table 4)**. In our analysis that evaluated the presence of NYHA class documentation in the year before ICD implantation, we found that among the 5,955 unique patient encounters, only 887 (14.9%) had an explicit mention of NYHA class in their clinical notes during that time period.

**Figure 6.**
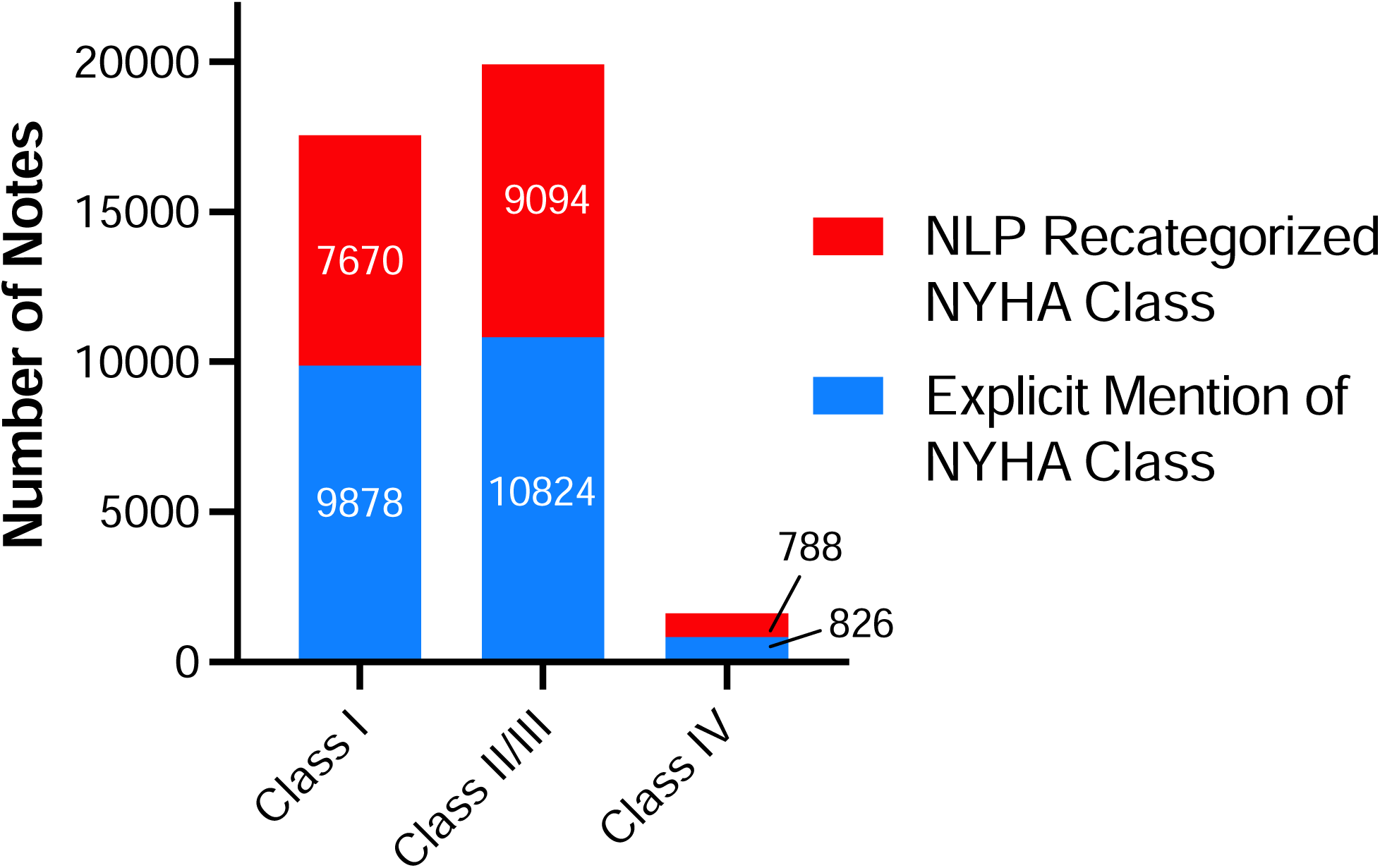
Frequency of Explicit and Natural Language Processing Recategorized New York Heart Association Class by Patient Note. Comparison of the number of explicit mentions in the notes to those reclassified through the NLP model across Classes I, II/III, and IV. Stacked bars represent the sum of notes for each NYHA class, highlighting the contribution of NLP in identifying potential cases that may have been otherwise undocumented. Abbreviations: NLP, Natural Language Processing; NYHA, New York Heart Association.

**Table 4.** Post-Deployment Assessment of Functional Status Across the Health System. The number of heart and vascular clinic outpatient encounter documentations that were categorized into each NYHA classes, among 166,655 records at YNHHS that were not used in either model development or validation, using the two NLP models developed in this study. Firstly, the NYHA class model was applied to identify explicit mention of NYHA class in the clinical documentations. Secondly, for the documentation without explicit mention of NYHA class, the symptom association model was employed to assign additional outpatient encounters into each NYHA classes, using the strategy illustrated in the Supplemental Figure 2. The two models categorized a total of 39,170 (23.5%) of all encounters into NYHA classes. Abbreviations: NYHA, New York Heart Association.

|  | Explicit NYHA Class Mention | Without Explicit Mention of NYHA Class | Recategorized NYHA Class | Combined NYHA Class after Recategorization |
| --- | --- | --- | --- | --- |
| NYHA Class I | 9,878 (5.9%) | 156,777 (94.1%) | 7,760 (4.6%) | 17,638 (10.5%) |
| NYHA Class II/III | 10,824 (6.5%) | 155,831 (93.5%) | 9,094 (5.4%) | 19,918 (11.9%) |
| NYHA Class IV | 826 (0.5%) | 165,829 (99.5%) | 788 (4.7%) | 1,614 (1.0%) |
| Total | 21,528 (12.9%) | 145,127 (87.1%) | 17,642 (10.5%) | 39,170 (23.5%) |

## DISCUSSION

In our study spanning multisite EHR data, we developed and validated a novel deep learning approach for unstructured medical notes to identify explicit mentions of NYHA functional status and to define activity-related symptoms and their classification into structured functional status groups when NYHA classes were not explicitly mentioned. Despite its critical role in therapeutic decisions, only 1 in 8 health encounters had any mention of NYHA classes in their notes. However, our work identified that the classification to an NYHA class based on text description could increase the capture of this information by 82%, to nearly 1 in 4 notes. The models exhibited consistently excellent performance across internal and external validation populations drawn from different types of practices, including a large academic hospital (YNHH), a community-based multispecialty group practice (NMG), and a community teaching hospital (GH), demonstrating the generalizability of our approach across diverse healthcare settings. The interpretable inference confirmed that the models learned textual signatures consistent with features used by human abstractors. This approach is able to identify gaps in clinical documentation while also providing a robust and scalable solution to track patient functional status and the alignment of treatment status with their functional status.

Our study builds upon prior efforts to extract functional status information from EHR data, offering a more efficient and scalable approach. Previous studies have employed rule-based algorithms or machine learning models to identify NYHA classes or functional status descriptors.^18,19^ However, these methods often struggle to capture the nuanced and varied ways in which functional status is documented in free-text narratives. In contrast, our deep learning NLP framework can process large volumes of unstructured data, identifying complex patterns and contextual information that may be missed by rules-based methods alone. This comprehensive approach enables a more accurate and complete assessment of a patient’s functional capacity across a wide range of clinical scenarios, which is essential for guiding HF management and ensuring adherence to best practices.

The implications of our findings are threefold. First, our NLP models can facilitate quality measurement initiatives by providing a reliable means to track functional status assessments and their impact on clinical decision-making. Second, the models can be integrated into clinical decision support systems, alerting providers when functional status assessments are due or when treatment plans may need adjustment based on a patient’s current functional capacity. This would also further increase the capture of this information in documentation via a clear identification of when this information has not been recorded. Finally, our approach can streamline patient identification for clinical trials by enabling the rapid screening of large patient cohorts based on their functional status, a key inclusion criterion for many HF studies.^20^

Interestingly, our interpretability analysis revealed that the models not only learned to recognize explicit mentions of NYHA classes but also picked up on key phrases and descriptors associated with different levels of functional impairment. This suggests that the models can capture the underlying clinical reasoning used by providers when assessing a patient’s functional status, even in the absence of standardized terminology. This capability is particularly valuable given the inherent variability in how functional status is documented across different providers and institutions.

Our study has certain limitations that merit consideration. First, the performance of our NLP models may be influenced by the specific annotation guidelines and criteria used during model development. While we aimed to create a generalizable framework, institution-specific documentation practices or variations in clinical terminology could impact the models’ accuracy when applied to other settings. Nevertheless, the various practice sites share few clinicians, are geographically separated, and have practice patterns largely governed by local patient populations, suggesting the likely generalizability of the tool outside the tested hospitals and clinics. Second, the complexity of HF symptoms and the potential for comorbid conditions to contribute to functional impairment may not be fully captured by our current models, particularly in more nuanced clinical scenarios. Future work should focus on refining the models to better account for these complexities and to incorporate additional clinical context when available. Finally, we opted for scalability and model efficiency over model size and, therefore, chose to use a ClinicalBERT model over more recently described large language models. However, given the high performance of our model in detecting the condition, along with the ease of deploying a model that does not require a large graphical processing unit capacity, we believe the decision to use a lightweight model that is easy to deploy in practice is appropriate.

## CONCLUSION

We developed and validated a deep-learning NLP approach to extract NYHA classification and activity-related HF symptoms from clinical notes. This scalable solution enables tracking whether patients receive optimal patient care and enhances the automated identification of those eligible for clinical trials using existing clinical documentation.

## Data Availability

The data cannot be publicly shared as it represents protected health information and sharing data will be a violation of patient privacy.

