## Supplement for "A Deep Learning Approach for Automated Extraction of Functional Status and New York Heart Association Class for Heart Failure Patients During Clinical Encounters"

**Supplementary Materials**

**Supplementary Methods - Annotating Guidelines**

The primary objective of our annotation task is to systematically classify notes and sentences based on HF functional status. At the note-level, patient notes were organized into categories of NYHA Class I, II, III, IV, or cases with no NYHA Class mentioned. This categorization helps establish the severity of heart failure in each patient, providing context for the functional status assessment. Each note is tokenized into sentences and initially labeled as either pertaining to 'functional status' or 'not functional status'. Subsequently, those identified as 'functional status' sentences undergo a more nuanced subclassification. These subclasses are designed to capture various aspects of a patient's functional status and include categories such as 'symptoms with activity', 'no symptoms with activity', 'symptoms with rest', 'no symptoms with rest', 'activity ability', and 'activity limitation'.

In the process of annotating outpatient medical notes, it is important for annotators to focus solely on the content within each sentence, without drawing upon the surrounding context. This approach ensures a standardized and objective analysis of each sentence, aligning with our goal of identifying HF functional status indicators. Annotators must operate under the assumption that any mention of symptoms or limitations in a patient's functional status is related to HF, except in cases where other specific causes are explicitly stated. For instance, a sentence stating "symptoms limit the patient's ability to walk up a flight of stairs" should be interpreted as HF-related unless other conditions are clearly identified.

Additionally, annotators are encouraged to consider the writer's intent and the clinical relevance of each sentence. Understanding why a healthcare provider chose to include specific information is crucial. For example, a sentence detailing a patient's daily activities or limitations provides valuable insight into their functional status and should be considered relevant to HF unless stated otherwise. This perspective aids in capturing the nuances of clinical communication, enhancing the quality and applicability of the annotated data. Annotators must strictly adhere to the provided definitions and guidelines, ensuring that each sentence is classified accurately and consistently.

**NYHA Class Manual Annotation**

The NYHA Classification labels are based on explicit mention of active current NYHA class categorization by a documented physician.

**Qualifying Criteria:**

- Sentences where a healthcare provider explicitly mentions active NYHA classification.
- Sentences describing symptoms clearly aligning with a specific NYHA class, even if the class is not explicitly mentioned.
- Direct statements about the patient's physical ability or limitations correlating with a specific NYHA class.

**Non-Qualifying Criteria:**

- Sentences that make broad statements about the patient’s health without specific reference to NYHA class or related symptoms.
- Descriptions of symptoms or limitations clearly attributed to conditions other than heart failure.
- Past or future medical history that does not reflect the current NYHA class.

**Notes and Edge Cases:**

- In cases where symptoms could be attributed to multiple classes, use the most restrictive class (highest number) that the symptoms could indicate.
- When a sentence contains both qualifying and non-qualifying information, prioritize the information that aligns with the NYHA classification.

**Symptom Association Manual Annotation**

The category of ‘Symptom Association’ encompasses sentences that explicitly describe a patient's physical abilities, limitations, or symptoms as they relate to daily activities and rest, particularly in the context of HF. To qualify as a ‘Symptom Association' sentence, the text must provide clear information about the patient's capability or incapacity to perform tasks, or the presence of symptoms during these activities or at rest.

**Qualifying Criteria:**

- Sentences that mention symptoms (like dyspnea, fatigue) occurring during physical activities.
- Sentences indicating symptoms experienced by the patient at rest.

**Non-Qualifying Criteria:**

- General health statements that do not specifically address the patient’s current functional status.
- References to medical treatments or medications without explicit mention of their impact on functional status.
- Historical information about the patient’s abilities or symptoms not relevant to their current condition.

**Notes and Edge Cases:**

In cases where the classification of a sentence is ambiguous or unclear, annotators should adhere to the following guidelines:

- When a sentence implicitly suggests a change in functional status without direct mention (e.g., "Patient now requires assistance for walking"), classify it as 'Functional Status'. The underlying assumption is that the change is significant enough to be noted in the patient's medical record.
- If a sentence describes symptoms without explicitly attributing them to HF but no other causes are mentioned (e.g., "Experiences shortness of breath while climbing stairs"), assume the symptoms are HF-related and classify accordingly.
- Distinguish between historical anecdotes and current functional status. Focus on the current state; historical references are not to be classified as 'Functional Status' unless they directly impact or explain the current condition.
- Broad or general statements about the patient’s health (e.g., "Patient is doing well") should not be classified under 'Functional Status' unless they include specific information about physical abilities or symptoms

**Supplemental Figure 1. Map of Internal and External Sites.** Each map pin represents an internal or external site of YNHHS where EHR data was collected, with blue representing Yale New Haven Hospital, orange representing Northeast Medical Group, and green representing Greenwich Hospital.

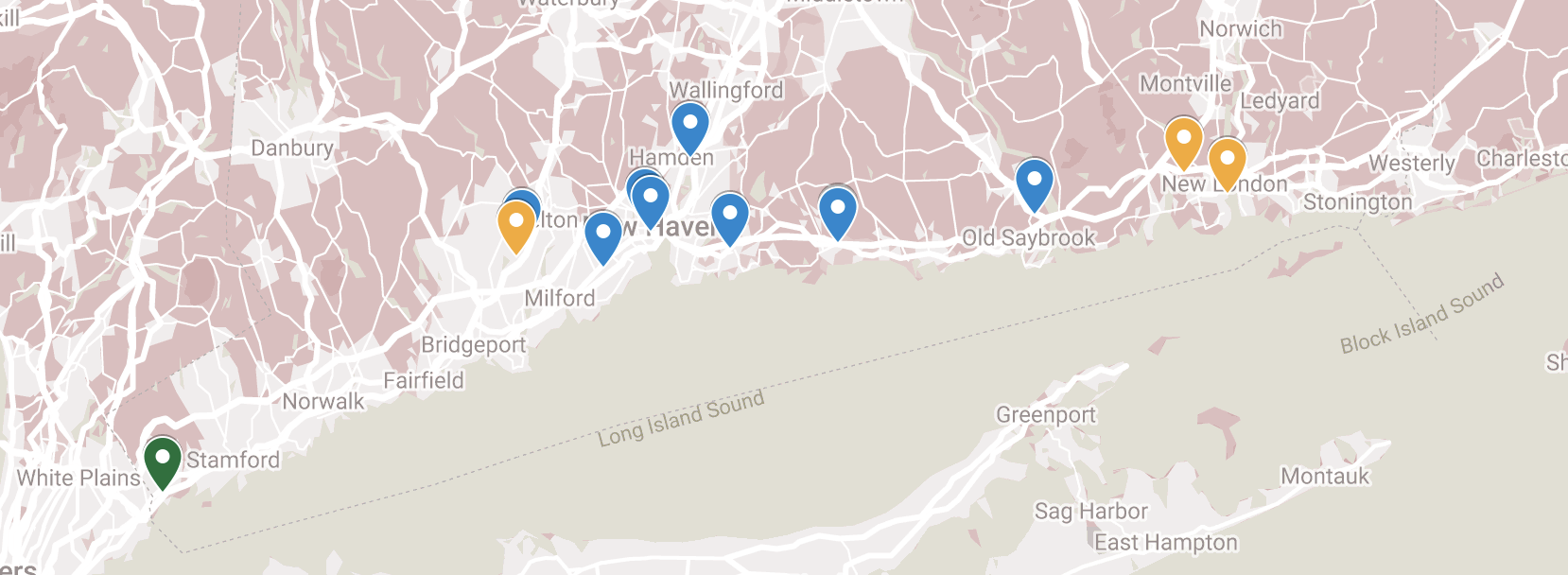

**Supplemental Figure 2. Algorithm for Recategorization.** Using the HF symptom association model, the strategy illustrated in this flowchart was employed to re-categorize patients without explicit mention of NYHA class in their clinical documentations to different NYHA classes

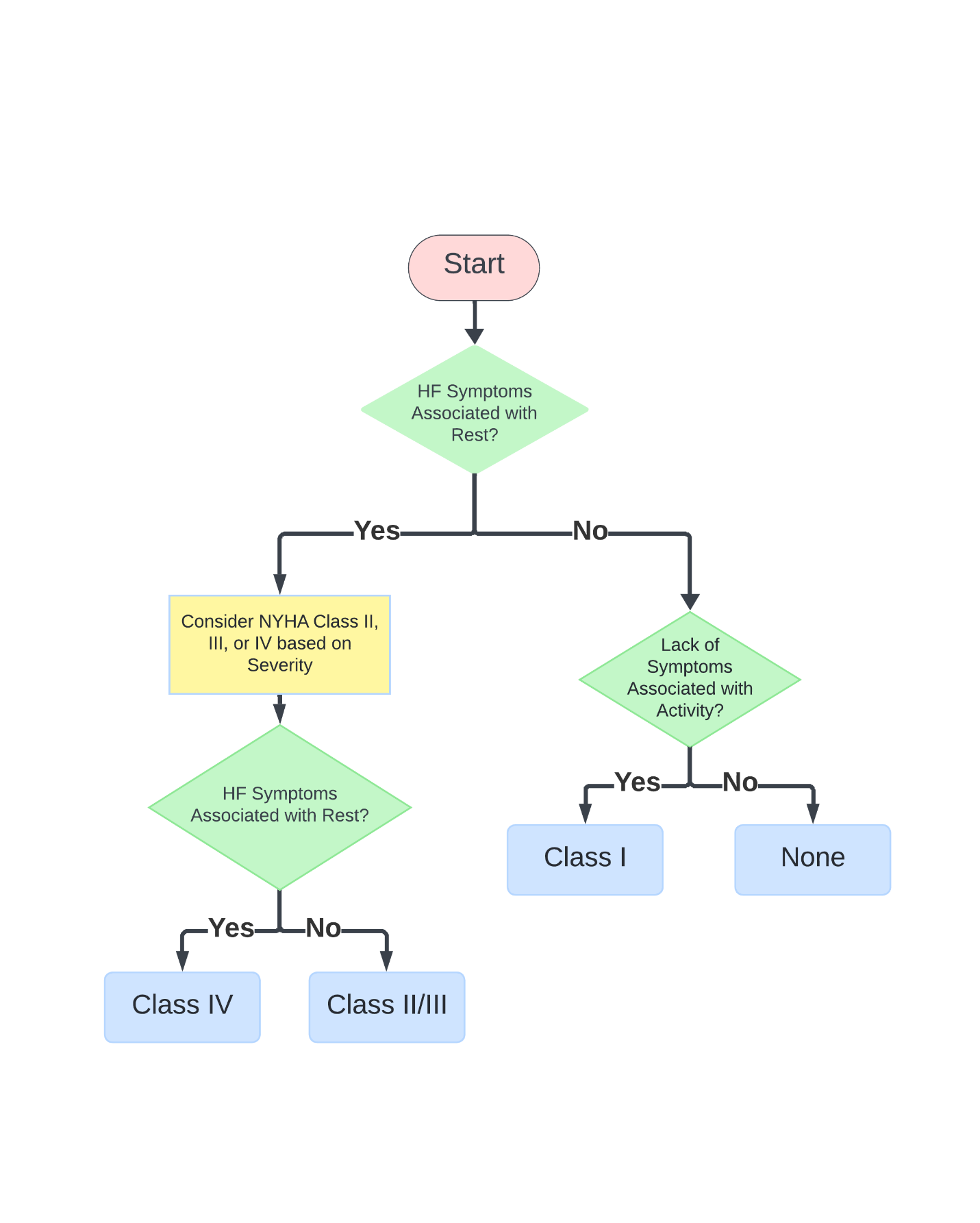

**Supplemental Table 1**. **Cardiovascular outpatient centers affiliated with Yale New Haven Hospital, Northeast Medical Group, and Greenwich Hospital that served as validation sites for the study.**

| Validation Site | Cardiovascular Outpatient Centers |
| --- | --- |
| Yale New Haven Hospital | Heart & Vascular Clinic North Haven, Heart & Vascular Clinic Branford, Heart & Vascular Clinic ORANGE', Heart & Vascular Clinic Guilford, Heart & Vascular Clinic Westbrook, Heart & Vascular Clinic Transition Care, Branford Ambulatory Cardiology Service, Yale New Haven Cardiac Rehab Center, Yale Medicine New Haven Sherman Avenue, Yale New Haven Congestive Heart Failure Clinic, New Haven Ambulatory Cardiology Service, Guilford Ambulatory Cardiology Service, Yale New Haven Prevention and Fellows Clinic, Old Saybrook Ambulatory Cardiology |
| Northeast Medical Group | Lawrence & Memorial Cardiology Waterford, Shelton Cardiology Group, New London Cardiology Group, Norwich Cardiology Group |
| Greenwich Hospital | Greenwich Hospital Outpatient Cardiology |

**Supplemental Table 2. International Classification of Disease Diagnosis Codes for Identification of Patients with Heart Failure and Comorbidities.** We identified heart failure patients based on ICD code as documented in the EHR at Yale New Haven Health System.

| Condition | ICD-9 Codes | ICD-10 Codes |
| --- | --- | --- |
| Heart Failure | 402.x, 404.x, 428.0, 428.1, 428.2, 428.20, 428.21, 428.22, 428.23, 428.3, 428.30, 428.31, 428.32, 428.33, 428.4, 428.40, 428.41, 428.42, 428.43, 428.9 | I50.1, I50.2, I50.20, I50.21, I50.22, I50.23, I50.3, I50.30, I50.31, I50.32, I50.33, I50.4, I50.40, I50.41, I50.42, I50.43, I50.8, I50.81, I50.810, I50.811, I50.812, I50.813, I50.814, I50.82, I50.83, I50.84, I50.89, I50.9 |
| Acute Myocardial Infarction | 410.x | I21.x, I22.x |
| Cardiomyopathy | 425.x | I42.x, I43.x |
| Hypertension | 401.x, 402.x, 403.x, 404.x, 405.x | I10, I11.x, I12.x, I13.x, I15.x |
| Diabetes | 250.x | E08.x, E09.x, E10.x, E11.x, E13.x |
| Chronic Kidney Disease | 585.x, 586 | N18.x, N19 |

**Supplemental Table 3. Dictionary and CPT Codes for Identifying ICD Implantation and Spironolactone Initiation**

| **Procedure/Medication** | **Dictionary/CPT Codes** |
| --- | --- |
| **ICD Implantation** | 'Insert of Defib Gen into Chest Subcu/Fascia, Open Approach', 'Insertion of Defib Lead into R Ventricle, Perc Approach', 'Insertion of Defibrillator Lead into R Atrium, Perc Approach', 'Insert Card Rsync Defib Puls Gen in Chest Subcu/Fascia, Open', 'Insertion of Defibrillator Lead into Cor Vein, Perc Approach', 'IMPLT/REPL CARDDEFIB TOT', 'IMPL CRT DEFIBRILLAT SYS', 'IMP/REP CRT DEFIB GENAT', 'Insertion of Defib Lead into L Ventricle, Perc Approach', 'Insertion of Defib Lead into Pericardium, Open Approach', 'Insertion of Subcutaneous Defibrillator Lead into Chest Subcutaneous Tissue and Fascia, Open Approach', 'Insertion of Subcutaneous Defibrillator Lead into Chest Subcutaneous Tissue and Fascia, Percutaneous Approach', 'Insert Card Rsync Defib Puls Gen in Chest Subcu/Fascia, Perc', 'Insert of Defib Gen into Chest Subcu/Fascia, Perc Approach', 'Insertion of Defib Lead into R Ventricle, Open Approach', 'Insertion of Defib Lead into L Ventricle, Open Approach', 'IMPLT CARDIODEFIB LEADS', 'Insertion of Defib Lead into Pericardium, Perc Approach', 'Insertion of Defib Gen into Abd Subcu/Fascia, Open Approach', 'Insert Card Rsync Defib Puls Gen in Abd Subcu/Fascia, Open', 'Insertion of Defibrillator Lead into L Atrium, Open Approach', 'Insertion of Defibrillator Lead into L Atrium, Perc Approach', 'Insertion of Defibrillator Lead into R Atrium, Open Approach', 'Insert Card Rsync Defib Puls Gen in Abd Subcu/Fascia, Perc', 'Insertion of Defibrillator Lead into Cor Vein, Open Approach', 'Insertion of Defib Lead into R Atrium, Perc Endo Approach', 'IMPLT CARDIODEFIB GENATR', 'Insertion of Defib Lead into L Ventricle, Perc Endo Approach', 'Insertion of Defib Lead into L Atrium, Perc Endo Approach', 'Insertion of Defib Lead into R Ventricle, Perc Endo Approach', 'Insertion of Defib Lead into Cor Vein, Perc Endo Approach', '33216', '33217', '33225', '33230', '33231', '33240', '33249', '33270', '33271', 'C7537', 'C7538', 'C7539', 'G0448' |

**Supplemental Table 4. Frequency of NYHA Classification in Manual Annotation of Outpatient Notes.** This table displays the count and proportion of outpatient notes manually annotated for NYHA Class I, II, III, IV, and notes with no NYHA classification at Yale New Haven Hospital, Northeast Medical Group, and Greenwich Hospital.

|  | NYHA Class I | NYHA Class II | NYHA Class III | NYHA Class IV | None |
| --- | --- | --- | --- | --- | --- |
| Yale New Haven Hospital (n = 2000) | 57 (2.9%) | 118 (5.9%) | 86 (4.3%) | 10 (0.5%) | 1729 (86.4%) |
| Northeast Medical Group (n = 500) | 12 (2.4%) | 40 (8.0%) | 26 (5.2%) | 2 (0.4%) | 420 (84.0%) |
| Greenwich Hospital (n = 500) | 8 (1.6%) | 9 (1.8%) | 4 (0.8%) | 2 (0.4%) | 477 (95.4%) |

**Supplemental Table 5. Documentation of Heart Failure Symptoms in Outpatient Notes.** Distribution of documented HF symptoms in outpatient notes at the three validation sites. Percentages reflect the proportion of notes within each category relative to the total number of notes reviewed at Yale New Haven Hospital, Northeast Medical Group, and Greenwich Hospital.

|  | | **Symptoms with Activity** | **Lack of Symptoms with Activity** | **Symptoms with Rest** | **Lack of Symptoms with Rest** | **None** |
| --- | --- | --- | --- | --- | --- | --- |
| **Yale New Haven Hospital** | Notes (n = 2000) | 486 (24.3%) | 329 (16.5%) | 45 (2.3%) | 53 (2.7%) | 1087 (54.4%) |
| **Northeast Medical Group** | Notes (n = 500) | 125 (25.0%) | 54 (10.8%) | 10 (2.0%) | 7 (1.4%) | 304 (60.8%) |
| **Greenwich Hospital** | Notes (n = 500) | 39 (7.8%) | 38 (7.6%) | 3 (0.6%) | 1 (0.2%) | 419 (83.8%) |

**Supplemental Table 6. Performance Metrics of NLP Models in Classifying Each Functional Status Label.** This table presents the performance of each class in either the NYHA class model or the symptom association model, measured by accuracy, precision, recall, specificity, AUROC, AUPRC, and F_1_-score. 95% confidence intervals were obtained from bootstrap sampling with 1,000 iterations.

|  | **Accuracy (95% CI)** | **Precision (95% CI)** | **Recall (95% CI)** | **Specificity (95% CI)** | **AUROC (95% CI)** | **AUPRC (95% CI)** | **F_1_-score (95% CI)** |
| --- | --- | --- | --- | --- | --- | --- | --- |
| **NYHA Class I** | 0.97 (0.96-0.98) | 0.22 (0.19-0.24) | 1.00 (1.00-1.00) | 0.97 (0.96-0.98) | 0.98 (0.93-1.00) | 0.22 (0.19-0.24) | 0.36 (0.33-0.38) |
| **NYHA Class II** | 0.98 (0.97-0.99) | 0.64 (0.61-0.67) | 0.97 (0.97-0.98) | 0.98 (0.97-0.99) | 0.98 (0.95-1.00) | 0.62 (0.60-0.65) | 0.77 (0.75-0.80) |
| **NYHA Class III** | 0.98 (0.97-0.99) | 0.57 (0.54-0.60) | 0.94 (0.92-0.95) | 0.98 (0.97-0.99) | 0.96 (0.91-1.00) | 0.53 (0.51-0.56) | 0.71 (0.68-0.73) |
| **NYHA Class IV** | 1.00 (1.00-1.00) | 1.00 (1.00-1.00) | 1.00 (1.00-1.00) | 1.00 (1.00-1.00) | 1.00 (1.00-1.00) | 1.00 (1.00-1.00) | 1.00 (1.00-1.00) |
| **Symptoms with Activity** | 0.93 (0.91-0.95) | 0.93 (0.91-0.95) | 0.96 (0.95-0.98) | 0.88 (0.85-0.90) | 0.98 (0.96-0.99) | 0.98 (0.97-1.00) | 0.95 (0.93-0.97) |
| **Lack of Symptoms with Activity** | 0.96 (0.94-0.97) | 0.87 (0.84-0.90) | 0.98 (0.96-0.99) | 0.95 (0.93-0.97) | 0.99 (0.99-1.00) | 0.98 (0.96-0.99) | 0.92 (0.90-0.94) |
| **Symptoms with Rest** | 0.95 (0.93-0.97) | 0.72 (0.68-0.76) | 0.49 (0.44-0.53) | 0.99 (0.97-1.00) | 0.94 (0.92-0.96) | 0.71 (0.67-0.75) | 0.94 (0.92-0.96) |
| **Lack of Symptoms with Rest** | 0.98 (0.97-0.99) | 0.93 (0.91-0.95) | 0.57 (0.52-0.61) | 1.00 (0.99-1.00) | 0.92 (0.90-0.95) | 0.66 (0.62-0.71) | 0.70 (0.66-0.74) |
